# Right hemispheric white matter hyperintensities improve the prediction of spatial neglect severity in acute stroke

**DOI:** 10.1101/2022.04.08.22273547

**Authors:** Lisa Röhrig, Christoph Sperber, Leonardo Bonilha, Christopher Rorden, Hans-Otto Karnath

## Abstract

White matter hyperintensities (WMH) are frequently observed in brain scans of elderly people. They are associated with an increased risk of stroke, cognitive decline, and dementia. However, it is unknown yet if measures of WMH provide information that improve the understanding of poststroke outcome compared to only state-of-the-art stereotaxic structural lesion data. We implemented high-dimensional machine learning models, based on support vector regression (SVR), to predict the severity of spatial neglect in 103 acute right hemispheric stroke patients. We found that (1) the additional information of right hemispheric voxel-based topographic WMH extent indeed yielded an improvement in predicting acute neglect severity (compared to the voxel-based stroke lesion map alone). (2) Periventricular WMH appeared more relevant for prediction than deep subcortical WMH. (3) Among different WMH measures, voxel-based maps as measures of topographic extent allowed more accurate predictions compared to the use of traditional ordinally assessed visual rating scales (Fazekas scale, Cardiovascular Health Study scale). In summary, topographic WMH appears to be a valuable clinical imaging biomarker for predicting the severity of cognitive deficits and bears great potential for rehabilitation guidance of acute stroke patients.

## Introduction

White matter hyperintensities (WMH), also called white matter lesions or leukoaraiosis, are frequently seen as hyperintense areas on T2-weighted brain magnetic resonance imaging (MRI) of predominantly elderly people (Manolio et al., 1994; for review, see Tang et al., 2021). While not fully understood, both vascular and non-vascular causes are involved (e.g., Schmidt et al., 2011). WMH of presumed vascular origin are seen to represent features of the cerebral small vessel disease (Wardlaw et al., 2013; Wardlaw, Smith, & Dichgans, 2013). Because WMH increase with age (Manolio et al., 1994; Wardlaw et al., 2015; Wei et al., 2019), they were initially thought to be a consequence of the normal ageing brain. Recently, WMH have received more attention, as they are associated with an increased risk of stroke (Debette & Markus, 2010), stroke recurrence (Andersen et al., 2017; Georgakis et al., 2019), mortality (Debette & Markus, 2010; Georgakis et al., 2019), disturbed mobility (Guttmann et al., 2000), cognitive decline (Qi et al., 2019; Valdés Hernández et al., 2013), and dementia (Debette & Markus, 2010; Georgakis et al., 2019). Moreover, WMH are reported to be specifically related to recovery of cognitive but not other poststroke impairments (Hawe et al., 2018; Khan et al., 2019). Studies also find associations of WMH extent with worse cognitive functions in Parkinson’s disease (Huang et al., 2020; Linortner et al., 2020; Liu et al., 2021), as well as severity and rate of progression of primary progressive aphasia (Meier et al., 2020; Odolil et al., 2020). Other research reveals a connection between WMH severity and performance of chronic aphasia following stroke (Basilakos et al., 2019; Wilmskoetter et al., 2019). In sum, WMH seem to be a strong indicator of compromised brain health and cognitive reserve.

Only a few studies have investigated whether the extent of WMH is also a predictor of the occurrence and severity of cognitive impairment in the acute phase of stroke. Bahrainwala and colleagues (2014) reported an association between WMH severity (adjusted for age, sex, race, and infarct volume) and the probability of having spatial neglect after an acute right hemispheric stroke. Stroke patients with spatial neglect show spontaneous and sustained deviation of their eyes and head as well as focus of attention toward the ipsilesional, typically right side of space (Karnath & Dieterich, 2006; Karnath & Rorden, 2012). Simultaneously, they fail to observe and react to stimuli, objects, and people on the contralesional side. Bahrainwala and colleagues (2014) found that for the same size of stroke lesion, a greater extent of WMH increased the risk of suffering from any and also more severe spatial neglect in the acute phase of stroke. Of high interest, the authors provided evidence that prestroke WMH affect poststroke acute attentional impairments. Such biomarkers could be used in the guidance and planning of poststroke rehabilitation and care.

Unfortunately, this seminal study used data with reduced information content as predictors, namely infarct volume without localization-related information and the extent of WMH as an ordinal scaled measure. However, voxel-wise representation is commonly used in state-of-the-art prediction algorithms (Hope et al., 2018; Salvalaggio et al., 2020; Siegel et al., 2016), and previous research demonstrated that voxel-based maps are better able to describe the brain’s complexity compared to lesion volume (Rondina et al., 2016) and are thus likely to achieve more accurate behavioral predictions. The ordinal clinical visual rating scales used for the WMH assessment bear also the risk of producing subjective discrepancies between raters. Moreover, Bahrainwala and colleagues (2014) predicted dichotomized variables, i.e., the presence versus absence of spatial neglect, despite the fact that symptom severity varies between patients (Rorden & Karnath, 2010). Furthermore, the authors did not test whether the localization of WMH has any significance for the severity of acute spatial neglect. Specifically, are left-sided, right-sided, and bilateral WMH equally predictive, and is it relevant whether the WMH are located near the lateral ventricles or within the deep subcortical white matter?

The first aim of the present study was to investigate whether the precise, voxel-based measurement of WMH extent by using semi-automatic MRI analysis improves the accuracy of predicting neglect severity in acute stroke patients, compared to using rating-based measures. We tested the visual rating scales by both the Cardiovascular Health Study (CHS; Manolio et al., 1994), which was applied by Bahrainwala and colleagues (2014), and Fazekas (Fazekas et al., 1987), which is the most commonly used WMH rating scale (Georgakis et al., 2019). Furthermore, instead of determining only the presence versus absence of spatial neglect, we analysed the deficit as a continuous variable. We thus applied machine learning-based support vector regression (SVR) as it allows to predict continuous variables with high-dimensional features. Second, we were interested in examining the possible impact of WMH localization within the hemisphere(s) on predicting the severity of neglect.

## Methods

### Participants

We included 103 individuals with right hemisphere stroke who were consecutively admitted to the Center of Neurology of Tübingen University; for demographic and clinical data, see Table 1. The patients had participated in previous studies of our group. Inclusion criteria for the current investigation were first-ever, right hemispheric stroke with a demarcated lesion and available MRI scans including a fluid-attenuated inversion recovery (FLAIR) image. Patients were not included when the period between stroke onset and imaging or neuropsychological examination was longer than 16 days. Patients with bilateral strokes or brain tumors were also not included. All patients gave their informed consent for participation and scientific data usage. The study was conducted in accordance with the revised Declaration of Helsinki and was approved by the ethics committee of the medical faculty of Tübingen University.

**Table 1.** Demographic and clinical data of the patient sample.

| Parameter | Total<br>(*N* = 103) | Neglect<br>(*N* = 27) | No neglect<br>(*N* = 76) | *p*-value |
| --- | --- | --- | --- | --- |
| Age (years) | 58.2 (13.7) [26–82] | 60.5 (15.1) [27–82] | 57.4 (13.2) [26–80] | 0.354<sup>a</sup> |
| Sex (*F*, *M*) | 37, 66 | 10, 17 | 27, 49 | 0.888<sup>b</sup> |
| Days between stroke–<br>imaging | 3.5 (3.9) [0–16] | 4.7 (4.7) [0–15] | 3.1 (3.5) [0–16] | 0.134<sup>a</sup> |
| Etiology (*I*, *H*) | 96, 7 | 26, 1 | 70, 6 | 0.457<sup>b</sup> |
| Lesion volume (cm<sup>3</sup>) | 27.5 (32.9) [0.1–182.0] | 55.6 (44.4) [0.4–182.0] | 17.5 (20.1) [0.1–98.9] | **< 0.001<sup>a</sup>** |
| Days between stroke–<br>assessment | 3.4 (2.9) [0–14] | 3.0 (2.3) [0–8] | 3.5 (3.0) [0–14] | 0.393<sup>a</sup> |
| Letter CoC | 0.10 (0.23) [-0.06–0.96] | 0.39 (0.30) [0.01–0.96] | 0.01 (0.02) [-0.06–0.07] | **< 0.001<sup>a</sup>** |
| Bells CoC | 0.13 (0.24) [-0.03–0.92] | 0.46 (0.27) [0.12–0.92] | 0.02 (0.03) [-0.03–0.14] | **< 0.001<sup>a</sup>** |
| Mean CoC | 0.12 (0.23) [-0.01–0.91] | 0.43 (0.28) [0.08–0.91] | 0.01 (0.02) [-0.01–0.06] | **< 0.001<sup>a</sup>** |
| Visual field defects (*N*) | 12 | 3 | 9 | 0.919<sup>b</sup> |

Patients with mean CoC values of at least 0.082 were defined as having spatial neglect (cut offs for the letter and bells cancellation tasks are 0.083 and 0.081, respectively [Rorden & Karnath, 2010]). Results are given in either mean (standard deviation) [range] or number of patients. Lesion volume represents the normalized volume in MNI space. *p*-values of unequal variances *t*-tests ('a', continuous measure) or Chi<sup>2</sup>-tests ('b', categorical measure) are displayed. *p*-values < 0.05 were considered significant and are highlighted in bold type. Abbreviations: *F* – females, *M* – males, *I* – Ischemia, *H* – Hemorrhage, CoC – Center of Cancellation (Rorden & Karnath, 2010), *N* – number of patients.

### Behavioral data

Patients were tested for spatial neglect using the letter cancellation test (Weintraub & Mesulam, 1985) and bells cancellation test (Gauthier et al., 1989) as standard paper-and-pencil tests. We calculated the *Center of Cancellation* (CoC; Rorden & Karnath, 2010) as a continuous score ranging between -1 (right-sided neglect) and +1 (left-sided neglect). The CoC is a sensitive measure capturing both the number of omissions as well as their location. For each patient, the mean CoC of both cancellation tasks was used for the analyses. If one of both test results was not available (*N* = 5), we used the remaining CoC score. Primary visual field defects (hemi- or quadrantanopia) were assessed via usual neurological confrontation testing.

### Imaging data and analysis

We used the axial MR images acquired during the patients’ clinical investigation at the Center of Neurology Tübingen. Since WMH are most visible on FLAIR scans (e.g., Wardlaw et al., 2015), we used these for the assessment of WMH extent. For quantification of the stroke lesion, we used co-registered diffusion-weighted imaging (DWI) scans if imaging was acquired within 48 hours after stroke onset (*N* = 47); otherwise, we used the FLAIR scans again (*N* = 56).

For lesion volume quantification, we applied the following steps performed by the use of SPM 12 (*Statistical Parametric Mapping*; Wellcome Department of Imaging Neuroscience, London, UK) for MATLAB R2019a (The MathWorks, Inc., Natick, USA). Stroke lesion delineation was conducted semi-automatically, using the ‘Clusterize Toolbox’ (Clas et al., 2012; de Haan et al., 2015) implemented in SPM. Clusterize is an algorithm automatically marking hyperintense (or hypointense) areas according to an intensity threshold; we used the default minimum cluster size of 100 voxels (Clas et al., 2012). A voxel-based binary brain map was generated by selecting the clusters belonging to the lesion for each image slice separately by one investigator (LR) who was blind to the behavioral measures. The resulting lesion map was then normalized (together with the respective anatomical image) to MNI space (*Montreal Neurological Institute*; Collins et al., 1994) with voxels of size 1 mm³ by using the ‘Clinical Toolbox’ (Rorden et al., 2012) for SPM and its age-matched templates (this step leveraged the high-resolution T1-weighted scan, if available). Lesions were controlled by either cost-function masking or enantiomorphic correction (cf. Karnath et al., 2019) depending on the available scan modalities and normalization outcome. Lateral ventricles, cerebellum and extracerebral space were masked to address normalization inaccuracies.

The extent of WMH was first determined quantitatively by directly obtaining each patient’s volumetric WMH map. For delineation and normalization of the WMH map, we used the same semi-automatic procedure via the ‘Clusterize Toolbox’ as for the patients’ volumetric stroke lesion map (see above), except that we now set the minimum cluster size to 10 voxels because WMH may consist of small punctate foci. WMH volume maps were normalized together with the same patient-specific spatial transformation parameters as for the patient’s stroke lesion normalization step. To exclude any potential overlap between both stroke and WMH maps in cases where WMH were adjacent to the stroke lesion, the WMH map was masked by the respective lesion map for each patient. For investigating laterality effects, we defined left and right WMH maps by separating the overall normalized WMH map at the midsagittal plane.

Second, the extent of WMH was also assessed visually by using two visual rating scales: (1) the scale described in the *Cardiovascular Health Study* (CHS; Manolio et al., 1994) applied by Bahrainwala et al. (2014) and (2) the more frequently used rating scale by Fazekas (Fazekas et al., 1987). Both scales are based on the examiner’s subjective visual inspection of the original FLAIR images. The CHS visual rating scale is based on 8 images depicting increasing severity stages of WMH; the scale ranges from 0 to 9, where 9 is defined as most severe. The Fazekas rating scale assesses periventricular white matter hyperintensities (PV-WMH) and deep subcortical white matter hyperintensities (DS-WMH) separately. Ratings are given as follows: PV-WMH: 0 – absent, 1 – ‘caps’ or pencil-thin lining, 2 – smooth ‘halo’, 3 – irregular PV-WMH extending into the deep white matter; DS-WMH: 0 – absent or a single punctate focus, 1 – multiple punctate foci, 2 – beginning confluency of foci, 3 – large confluent areas. The total Fazekas score is calculated by summing up PV-WMH and DS-WMH scores and therefore ranges from 0 to 6, where 6 is defined as most severe. One author (LR) conducted the ratings of all patients that were used in the analyses without knowing any clinical or neuropsychological data and diagnoses. To check for interrater reliability, another author (HOK) rated a subset of 20 examples; again without knowing any clinical or neuropsychological data and diagnoses. For each rating scale, we calculated Cohen’s linearly weighted kappa (*κ*) with the relative agreement to estimate consensus and Spearman correlation (*r*_*s*_) to estimate consistency: CHS scale – *κ* = 0.72 (90.7%) and *r*_*s*_ = 0.87; Fazekas total score – *κ* = 0.40 (82.5%) and *r*_*s*_ = 0.85; Fazekas PV-WMH score – *κ* = 0.58 (88.3%) and *r*_*s*_ = 0.76; Fazekas DS-WMH score – *κ* = 0.25 (73.3%) and *r*_*s*_ = 0.71. Although the weighted kappa for the DS-WMH score represents only fair consensus in rating (Landis & Koch, 1977; Stemler, 2004), the corresponding strong correlation demonstrates high consistency, which is sufficient for a reliable regression analysis.

### Prediction of spatial neglect severity

To investigate whether WMH volume information – in addition to the acute stroke lesion – increases prediction accuracy of the severity of spatial neglect, we applied the supervised learning algorithm support vector regression (SVR; Brereton & Lloyd, 2010; Smola & Schölkopf, 2004) that can predict a continuous variable. SVR was implemented using the ‘libsvm’ package (Chang & Lin, 2011) for MATLAB via custom scripts. The feature matrix consisted of the lesion map and – in the case of the combined models (see below for details) – the WMH data of all patients. Matrix rows comprised patients, while columns contained features of the predictive model: vectorized volume map(s) including the dichotomized status (damaged or not) of all voxels, and optionally also the WMH rating score. The vector of the mean CoC-scores, reflecting the severity of spatial neglect, served as the dependent variable. Since for our sample of only right hemispheric stroke patients no right-sided neglect was expected, negative mean CoC-values (*N* = 13 with a *mean original CoC* = -0.006) were set to 0 to avoid difficulties in interpreting the directionality of predicting a bi-polar target variable. Moreover, we square root transformed the mean CoC-values to de-skew our data as implemented in previous research (e.g., Kasties et al., 2021).

In a pilot investigation, we found worse model performances when the ordinal WMH ratings were transformed into dummy variables. We therefore treated the WMH scores as continuous measures. We applied the mean normalization for the WMH scale values to obtain features between 0 and 1 (i.e., the highest WMH rating score was assigned to the value of 1, whereas the smallest WMH rating score was assigned to the value of 0). To focus only on the most relevant voxels of the lesion maps, we excluded voxels that were damaged in less than 5 patients in all models investigated. In contrast, we altered this criterion for the inclusion of voxels in the WMH maps. Ordinal ratings took the wholistic WMH pattern into account, including brain regions that were only rarely affected by WMH, such as small foci of DS-WMH. Since we aimed to directly compare rating-based with voxel-based WMH measures, we included all WMH map voxels that were damaged at least once to make rating-based and topographic data comparable. In addition, we applied a principal component analysis (PCA) to reduce the high-dimensional feature space. This feature reduction procedure was shown to work best for high-dimensional data by yielding most accurate model fits (Kasties et al., 2021). We kept principal components that cumulatively explained at least 98% variance; these components were also normalized between 0 and 1. For feature reduction, the PCA was only applied to voxel-based matrices (lesion map, WMH map) without the inclusion of single-value variables (WMH scale ratings).

We implemented an epsilon-SVR with a radial basis function (RBF) kernel, where the default value for epsilon (*ε* = 0.1) was used in accordance with previous lesion-behavior modelling studies (e.g., Wiesen et al., 2019; Zhang et al., 2014). Previous studies investigating lesion-behavior relationships have reported better performance of nonlinear kernels compared to linear ones (Hope et al., 2018; Zhang et al., 2014). To avoid overfitting and to promote generalization of the fitted model, we implemented a nested cross-validation (CV) procedure (Fig. 1; Krstajic et al., 2014; Varoquaux et al., 2017). In the outer loop, we performed a 10-fold CV. Folds were almost equally sized. In one iteration, nine folds of the patient sample were assigned to the training-set and the remaining 10th-fold served as a test-set. The training-set of the outer loop was passed to the inner loop and contained all training- and validation-folds of the inner CV. In the inner loop, we had a 5-fold CV, i.e., four folds served as training-set of the inner CV and the remaining 5th-fold represented the validation-set. During the inner loop, the hyperparameters *C* and *γ* were optimized. For that, a grid search was implemented (*C* = 2^(−5)^, 2^(−4)^, …, 2^15^, and *γ* = 2^(−15)^, 2^(−14)^, …, 2^5^). Each combination of *C* and *γ* was trained on the training-sample and tested on the remaining validation-fold. We defined the *Mean Absolute Error* (MAE) as our parameter of interest. After the 5-fold CV (inner loop), the aforesaid parameter was averaged for that specific *C*-*γ*-combination. This procedure was repeated for each parameter combination. The algorithm was designed to minimize the MAE: the *C*-*γ*-combination that yielded the smallest mean MAE was considered to be the best model generated during the inner loop. The model with the winning hyperparameter-combination was then re-trained on the whole training-set of the outer loop and tested on the hold-out test-set. At the end of the whole nested CV, the outcome of each patient was predicted once during the outer loop.

**Figure 1.**
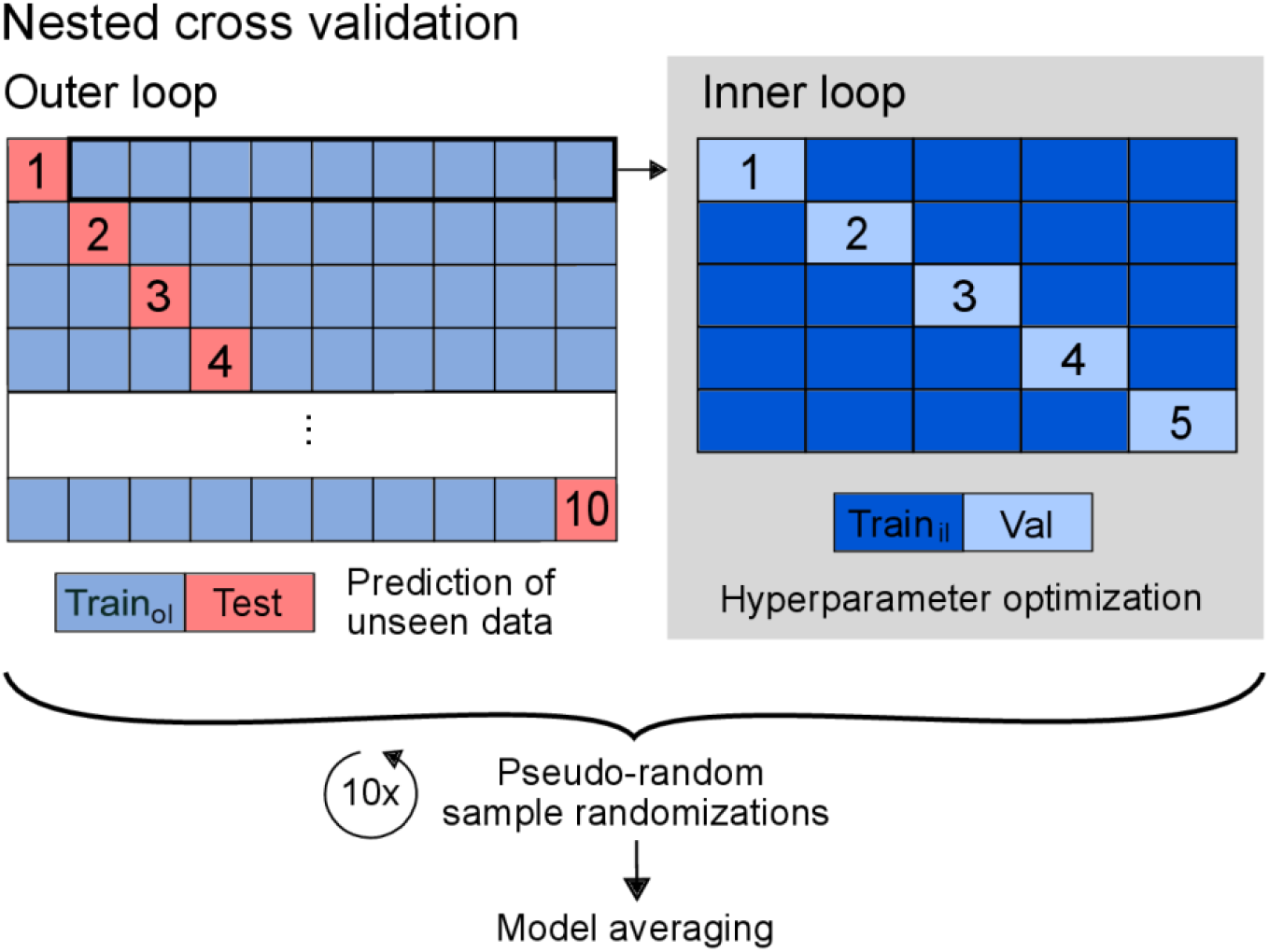
Overview of the applied nested cross-validation procedure. Nested cross-validation with 10-fold outer loop and 5-fold inner loop is illustrated. In the outer loop, 9 folds served as training set (‘Train_ol_’), which is passed to the inner loop. In the inner loop, 4 folds served as training set (‘Train_il_’), with which the hyperparameters *C* and *γ* were trained and tested on the remaining 5^th^ fold (validation set, ‘Val’). This optimization process aimed to find the model that minimizes the mean absolute error (MAE). The model with the winning *C*-*γ*-combination of the inner loop was then trained on the whole training set of the outer loop and tested on the hold-out 10^th^ test-fold. The model fit was calculated based on the predictions made during the outer loop. We used a pseudo-random number stream to shuffle the sample order and repeated the whole algorithm 10 times with different sample randomizations to include variance-driven effects. In the end, predictions of the 10 model repetitions were averaged to get the averaged model and to calculate the final model fit.

Since the resulting model fit is dependent on the patient assignments to training-, validation- and test-sets, we aimed to overcome that issue of variance by implementing model averaging (Arlot & Celisse, 2010; Varoquaux et al., 2017). To get a more generalized model fit, we repeated the whole model fitting procedure 10 times (cf. Fig. 1). For each model repetition, we applied a different sample randomization. For reproducibility and comparability issues, we used a specified random number stream. The sample was pseudo-randomized for each model repetition according to a sub-stream of the applied random number generator, so that the randomization of patients of a particular model repetition was the same across model versions. The predictions of all 10 model repetitions were then averaged for each patient. These final predictions were used to calculate the overall *Coefficient of Determination* (R²; for formula, see, e.g., Chicco et al., 2021), which represents the proportion of variance explained. The R² served as the model fit parameter when comparing the investigated SVR model versions as recommended by Chicco et al. (2021).

The feature matrix of the base model consisted of the voxel-based vectorized binary lesion map. For the other SVR models, we had to connect different types of data into one feature matrix. We used two methods of combining lesion and WMH: concatenation and addition of matrices (Fig. 2). (1) We concatenated the matrix of lesion maps and the vector of WMH scale values to get one enlarged matrix, separately for the CHS scale, Fazekas PV-WMH score, Fazekas DS-WMH score, and for both PV-WMH and DS-WMH scores concatenated. (2) We also merged the lesion matrix and the matrix of WMH maps into one concatenated large matrix to directly compare the effect of using volumetric information instead of a single rating. (3) Further, we performed a voxel-wise matrix addition to join the lesion and WMH maps together to one voxel-based volume. (For that, we kept for both volumetric maps all voxels that are lesioned at least 5 times among the lesion maps and/or at least 1 time among the WMH maps. The reduced lesion and WMH maps were added afterwards.) For the investigation of laterality effects, we also calculated models with either a left or a right hemispheric WMH map; here, we used the matrix fusion technique (matrix concatenation or addition) that revealed the best model performance for using the bilateral WMH map.

**Figure 2.**
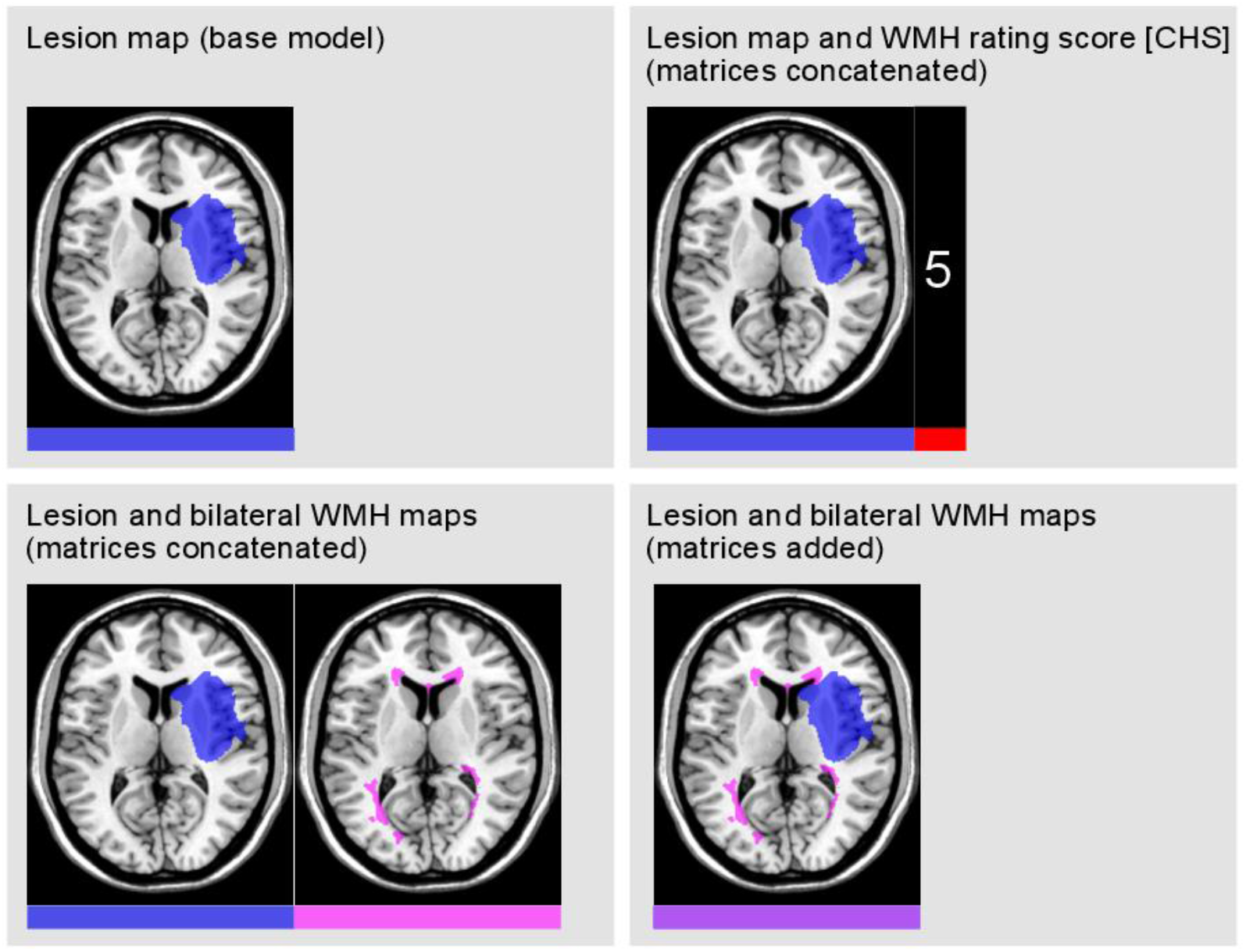
Illustration of the tested model versions and their feature matrices. Depending on the model, lesion map (blue) and white matter hyperintensities (WMH) map (pink) of an exemplary patient are presented on the ch2-template via MRIcron (Rorden & Brett, 2000). These matrices of damaged voxels were either attached to each other (concatenated) or matrices were summed up (added). In case of concatenation of lesion and WMH maps, we treated both variants of voxel-information as separate features. The matrix addition approach can be roughly explained as the simultaneous delineation of lesion and WMH in an overall brain damage map, as the expansion of the lesion map by the extent of WMH, or as the treatment of WMH as part of the brain lesion itself. For the WMH rating (CHS-scale exemplary), a single value representing the WMH severity was attached to the vectorized matrix of lesioned voxels (concatenated). When concatenating both periventricular and deep subcortical WMH ratings (Fazekas scale), two values were appended. Color bars illustrate the corresponding feature matrix (for one patient) that differed among the model versions.

We used principal components of the voxel-based brain maps as features for the prediction models. PCA revealed the numbers of components that are needed to cumulatively explain at least 98% variance: for lesion map 63 (out of 102), for concatenated matrices of lesion and bilateral WMH maps 88, for added matrices of lesion and bilateral/left/right WMH maps 88/83/84 components, respectively. An exploratory PCA of the bilateral WMH map alone resulted in 91 required components, indicating that WMH maps were more high-dimensional than lesion maps.

### Permutation tests

To validate whether a numerical increase in model fit is significantly above chance, we applied nonparametric Monte-Carlo simulation (permutation) testing to all models that yielded a better performance compared to the base model (the model that used only lesion data). For that, the model was calculated with a shuffled patient assignment of WMH data to investigate whether the patient-specific lesion-WMH-combination is relevant for obtaining an improved prediction performance. This way, we checked if WMH extent improves prediction significantly. In case of matrix addition, we controlled for any overlaps of lesion and WMH maps that may have occurred by setting all non-zeros to ones (keeping matrix binary). For each of the 10 model repetitions with different patient randomizations, we applied 5000 permutation iterations. The predictions of all 5000 permutations were averaged across the 10 model repetitions. For each of the 5000 averaged models with permuted WMH severity, the *Coefficient of Determination* (R²) was then calculated. From the distribution of 5000 R² obtained by the permuted variable (i.e., WMH data), one can conclude if the true WMH data resulted in a model performance significantly better than chance: if the 95%-best R² is smaller than the true R², then the true R² is significant on an alpha-level of 0.05. Using the predicted and true outcome scores, the p-value was calculated additionally. Since we were only interested in an improvement of prediction accuracy, we used a one-tailed significance test.

Again, for reproducibility issues, the random number stream that was implemented for the cross-validation procedure was also applied for the permutation process (5000 sub-streams were used to permute the WMH data pseudo-randomly). The use of pseudo-random permutations had the further advantage that we were able to directly compare the permutation tests of different model versions and their repetitions and to convincingly average the predictions that resulted from the 10 permutation repetitions.

## Results

### Clinical and anatomical aspects

Figure 3 illustrates the topography of simple overlaps of acute lesions and WMH in the group of right hemispheric stroke patients with and without spatial neglect (*N* = 27 and *N* = 76, respectively); Table 2 gives the results of the different measures of WMH for the two groups. A first result was that our continuously admitted sample of 103 acute stroke patients did not include patients without any WMH (i.e., CHS score of 0, WMH volume of 0 ml). With respect to the extent of WMH (regardless of whether measured ordinally or volumetrically), patients suffering from spatial neglect did not differ significantly from patients without the disorder (*p* > 0.05, Tab. 2). However, we observed numerically larger WMH volumes in patients with spatial neglect (cf. Tab. 2). Spearman correlations revealed highly significant positive relationships between all investigated WMH measures (*p* < 0.001, Tab. 3). The three variables CHS score, total Fazekas score and overall WMH volume (single value) were very strongly correlated (*r*_*s*_ ≥ 0.84; cf. Tab. 3); we thus expected similar results for SVR models using one of these as a predictor and therefore only investigated SVR models using the CHS rating. Moreover, we observed the previously described moderate positive relationships between age and the different measures of WMH extent (Tab. 3); no such correlations were found between these measures of WMH extent and neglect severity (*p* > 0.05).

**Figure 3.**
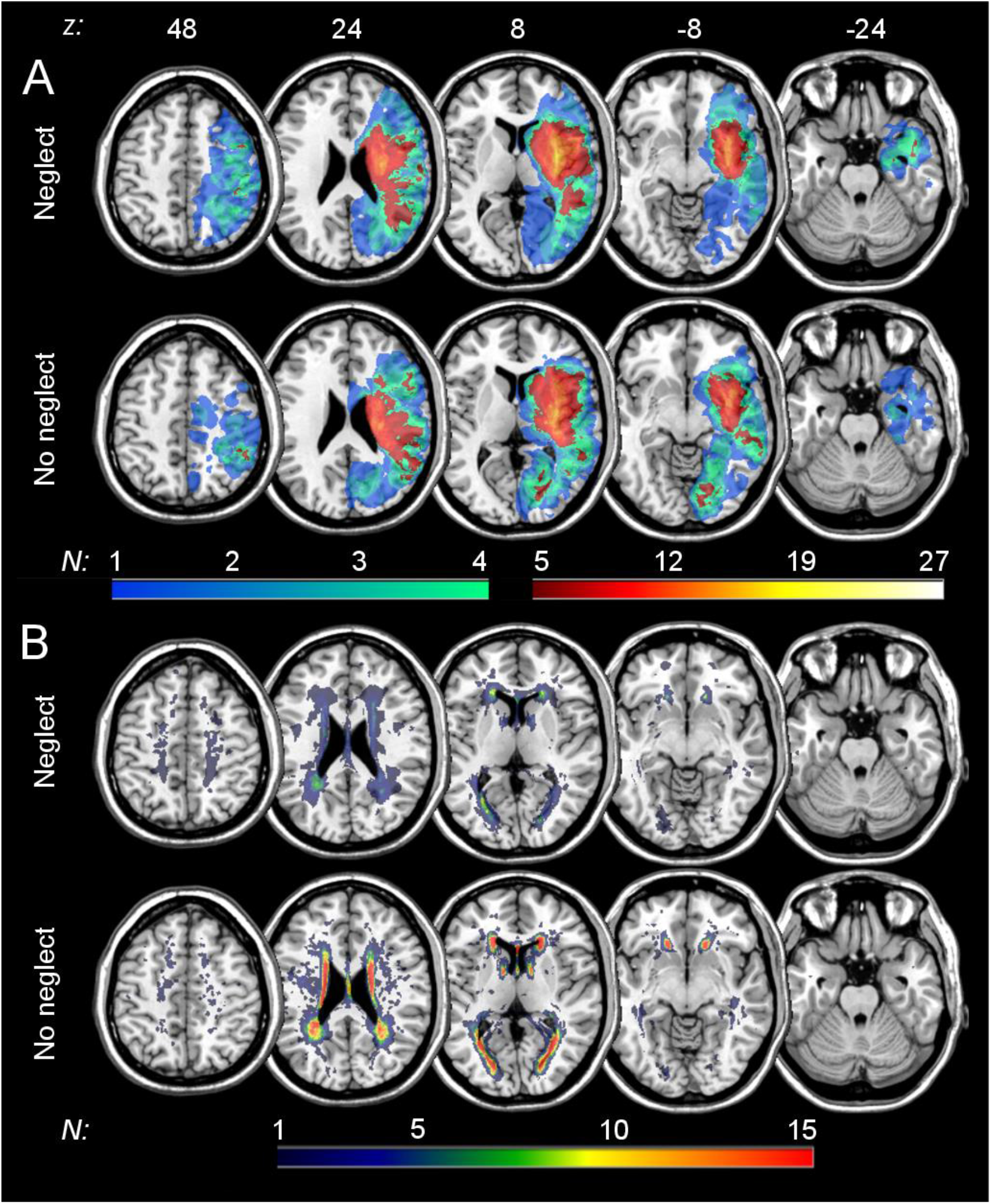
Simple lesion and WMH overlay plots of patients with and without spatial neglect. Overlaps of (**A**) normalized acute lesions and (**B**) normalized white matter hyperintensities (WMH) are shown for patients with and without spatial neglect (*N* = 27 or *N* = 76, respectively) on the ch2-template in MNI space via MRIcron (Rorden & Brett, 2000). Displayed axial slices refer to z-coordinates 48, 24, 8, -8 and -24 mm. The color of the voxels represents the number of patients with damage to this voxel (*N*_*min*_ = 1; *N*_*max*_ = 27 or *N*_*max*_ = 15, respectively). For lesions (**A**), cold colors refer to voxels that were damaged in less than 5 patients, whereas voxels displayed in warm colors (lesioned in at least 5 patients) were used for the support vector regression (SVR) analyses. For WMH (**B**), all voxels damaged in at least one patient were used for the SVR models.

**Table 2.** Different measures of white matter hyperintensities (WMH) extent for neglect versus non-neglect patients.

| Parameter | Total<br>(N = 103) | Neglect<br>(N = 27) | No neglect<br>(N = 76) | p-value |
| --- | --- | --- | --- | --- |
| CHS (0–9) | 3 [1–9] | 3 [1–9] | 3 [1–8] | 0.944<sup>a</sup> |
| Fazekas (0–6) | 2 [1–6] | 2 [1–6] | 2 [1–6] | 0.412<sup>a</sup> |
| PV-WMH (0–3) | 1 [0–3] | 1 [1–3] | 1 [0–3] | 0.901<sup>a</sup> |
| DS-WMH (0–3) | 1 [0–3] | 1 [0–3] | 1 [0–3] | 0.166<sup>a</sup> |
| WMH volume (cm<sup>3</sup>) | 8.0 (8.9) [0.4–64.8] | 9.6 (13.0) [1.2–64.8] | 7.5 (6.9) [0.4–39.4] | 0.432<sup>b</sup> |

Results are given as median [range] for ordinal data (for total scale range see parentheses) or mean (standard deviation) [range]. WMH volume represents the normalized volume in MNI space. *p*-values of Mann-Whitney U-tests ('a', ordinal measure) and unequal variances t-test ('b', continuous measure) are displayed. *p*-values < 0.05 were considered significant. Abbreviations: *CHS* – score of the Cardiovascular Health Study scale, *Fazekas* – total score of the Fazekas scale (scores of PV-WMH and DS-WMH summed up), *PV-WMH* – periventricular WMH score, *DS-WMH* – deep subcortical WMH score, *N* – number of patients.

**Table 3.** Correlations between patients’ clinical and demographic parameters.

|  |  |  |  |  |  |  |
| --- | --- | --- | --- | --- | --- | --- |
|  |  |  |  |  |  | CoC |
|  |  |  |  |  | Age | 0.16 |
|  |  |  |  | DS-WMH | 0.41 | 0.03 |
|  |  |  | PV-WMH | 0.36 | 0.39 | -0.02 |
|  |  | Fazekas | 0.74 | 0.87 | 0.48 | 0.01 |
|  | CHS | 0.86 | 0.67 | 0.74 | 0.53 | -0.03 |
| Volume | 0.89 | 0.84 | 0.61 | 0.75 | 0.45 | -0.08 |

Correlations were calculated between the different measures of white matter hyperintensities (WMH) extent, age and severity of spatial neglect (CoC score [Rorden & Karnath, 2010]). Correlation coefficients according to Spearman ( $r_s$ ), if an ordinal variable was involved, or Pearson ( $r_p$ ) are presented. All correlations were highly significant ( $p < 0.001$ ), except for the correlations involving the CoC score ( $p > 0.05$ ). Abbreviations: *Volume* – normalized bilateral WMH volume (in $\text{cm}^3$ ), *CHS* – Cardiovascular Health Study scale value, *Fazekas* – overall score of Fazekas scale (PV-WMH and DS-WMH scores summed up), *PV-WMH* – Fazekas periventricular WMH score, *DS-WMH* – Fazekas deep subcortical WMH score, *CoC* – Center of Cancellation.

### Prediction of spatial neglect severity

Figure 4 gives an overview of support vector regression (SVR) model performances, i.e., prediction accuracy of the averaged models as well as the performances of single model repetition iterations for all model versions; Figures S1 and S2 show corresponding scatter plots (supplementary material). The averaged base model that used only the lesion map (no additional WMH data) in the feature matrix revealed a prediction accuracy of *R²* = 0.331. Looking at performances of the single model repetitions (Fig. 4), the base model and the models using lesion map and WMH scale values (either CHS rating or Fazekas scores) showed variance in prediction accuracy depending on the sample randomization (*R²*_*max*_ – *R²*_*min*_ ≥ 0.102). Compared to that, models that used lesion and WMH maps (matrices either concatenated or added) seemed to be more stable in model fit across repetitions (*R²*_*max*_ – *R²*_*min*_ ≤ 0.079).

**Figure 4.**
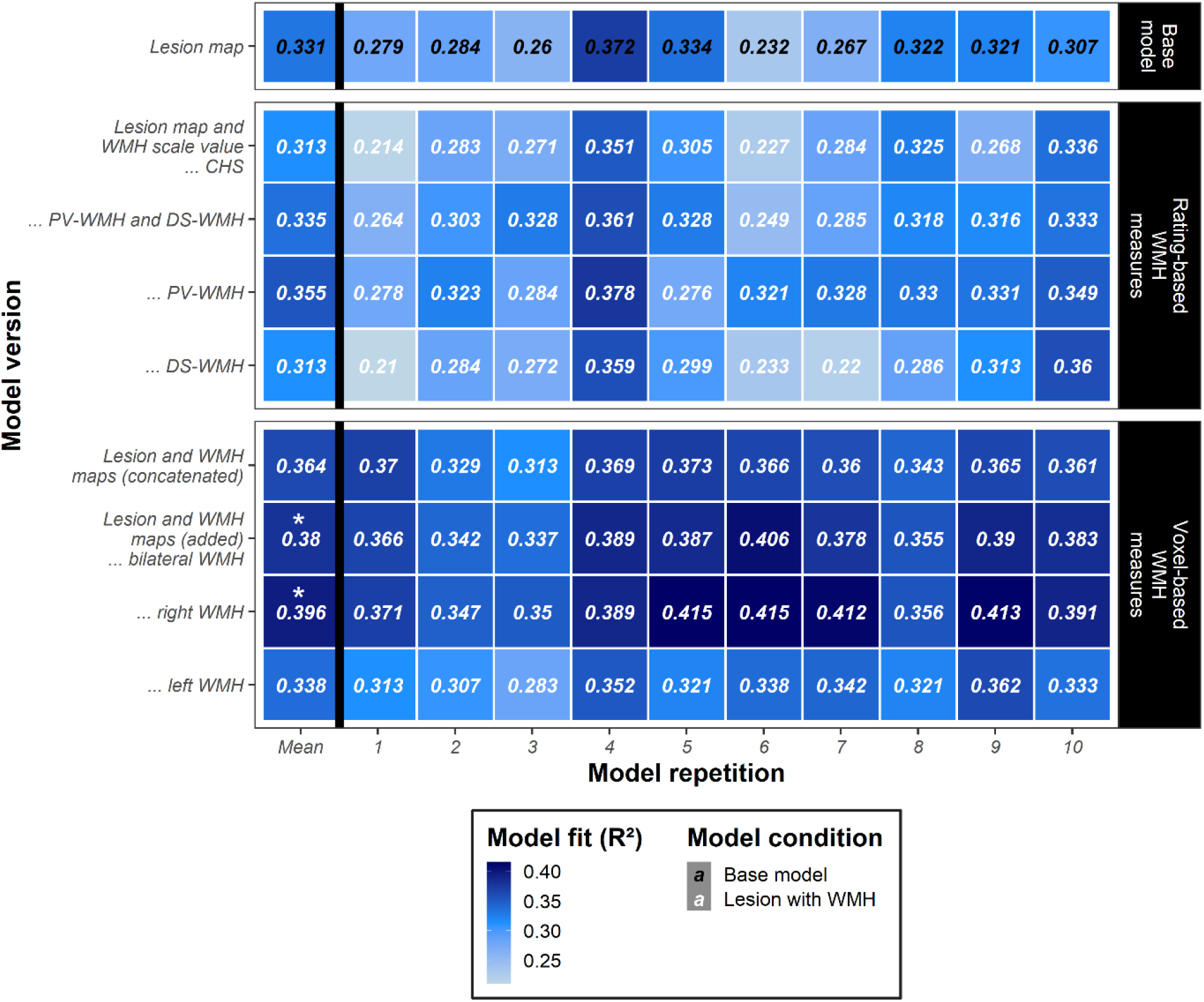
Prediction accuracies across model versions and repetitions. Heatmap illustrating accuracy of predicting neglect severity (CoC score [Rorden & Karnath, 2010]) for the cross-validated support vector regression (SVR) models. Prediction accuracy was measured as the *Coefficient of Determination* (R²) that represents the proportion of variance explained. The model versions differed in their feature matrix: the base model used only the lesion map for prediction (black font, upper segment), whereas the other models used in addition the extent of white matter hyperintensities (WMH, white font) as either ordinal visual scale ratings (middle segment; *CHS* – Cardiovascular Health Study, *PV-WMH* – periventricular WMH, *DS-WMH* – deep subcortical WMH, *PV-WMH and DS-WMH* – both vectors concatenated) or voxel-based maps (lower segment) as predictors. For more details on model versions, see Figure 2. For each model version, prediction accuracy is depicted for all 10 repetitions using different sample randomizations and for the averaged model (‘Mean’, predictions by model repetitions were averaged). Background tile color visualizes model performance, ranging from light blue (*R²* ≈ 0.2) to dark blue (*R²* ≈ 0.4). Averaged models that yielded a numerically improved model fit than the averaged base model were tested for significance by permutation testing; averaged models are highlighted with an asterisk if the original model using true WMH data showed a significantly improved model performance (*p* < 0.05; see Fig. 5).

#### Rating-based WMH measures

When attaching the CHS scale values concerning WMH extent to the lesion map matrix, the averaged model yielded a worse model fit of *R²* = 0.313. When using Fazekas scores instead, periventricular WMH (PV-WMH) and deep subcortical WMH (DS-WMH) yielded averaged model performances of *R²* = 0.355 and *R²* = 0.313, respectively (Fig. S2). The concatenation of both PV-WMH and DS-WMH scores resulted in *R²* = 0.335. For the scale ratings, the model using lesion map and PV-WMH score achieved the best model performance, indicating that PV-WMH was more predictive than DS-WMH. However, permutation testing revealed no improvement of prediction accuracy above chance (*p* > 0.05).

#### Voxel-based WMH measures

In comparison to ordinal WMH scale ratings, models that used volumetric WMH maps resulted in numerically better prediction accuracies (except for the model implementing the left hemispheric WMH map), suggesting that voxel-based WMH maps were more predictive than rating-based WMH measures. The concatenation of lesion and WMH maps resulted in minimum 0.9% and maximum 5.1% more explained variance compared to the concatenation of the lesion map and a visual scale rating. The concatenation of lesion map and bilateral WMH map resulted in an accuracy of *R²* = 0.364; however, permutation testing revealed no improvement of prediction accuracy above chance (*p* > 0.05). Opposed to that, matrix addition of lesion map and bilateral WMH map yielded a significantly improved prediction accuracy of *R²* = 0.380 (*p* = 0.026, Fig. 5A), indicating that the matrix addition resulted in more accurate models than the matrix concatenation. When using unilateral volumetric WMH maps for matrix addition (Fig. S2), no improvement of prediction accuracy above chance was found for the left hemispheric WMH map (*R²* = 0.338, *p* > 0.05). In contrast, results demonstrated a significantly better model performance than chance for the right hemispheric WMH map (*R²* = 0.396, *p* = 0.014, Fig. 5B). The achieved model fit for the additional right hemispheric WMH map was even larger than for the additional bilateral WMH map (cf. Fig. 4 and Fig. 5), suggesting that right hemispheric WMH were more predictive than left hemispheric (or bilateral) WMH. In sum, compared to the base model, the proportion of explained variance improved already by approximately 4.9% when adding the lesion map and the bilateral hemispheric WMH map together. The performance could even be further improved to approximately 6.5% increase in accuracy when adding the lesion map and the right hemispheric WMH map together. The latter was shown to be the overall best performing model, which explained nearly 40% of the total variance.

**Figure 5.**
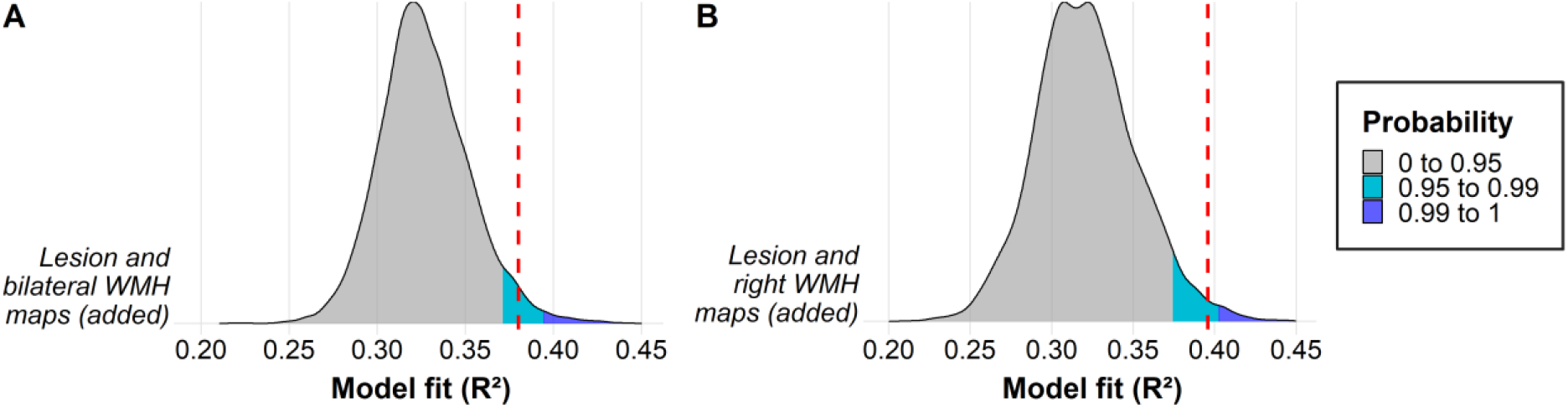
Permutation tests revealing measures of white matter hyperintensities (WMH) extent important for prediction. Distribution plots are shown for Monte-Carlo simulation (permutation) tests of 5000 iterations (predictions of permutation models were averaged across 10 model repetitions). The extent of WMH served as the permuted variable. Prediction accuracy was measured as the *Coefficient of Determination* (R²) that represents the proportion of variance explained. R²-distributions are displayed for the two model versions, for which the averaged model that used the true WMH data yielded a significant improvement of prediction accuracy above chance: addition of voxel-based volumetric maps containing (**A**) lesion and bilateral WMH and (**B**) lesion and right hemispheric WMH, indicating that bilateral and right hemispheric WMH maps were important features for prediction. The achieved model fit of the corresponding original model is visualized as a red dashed line (lesion and bilateral WMH: *R²* = 0.380; lesion and right hemispheric WMH: *R²* = 0.396). Areas of R²-distribution that represent one-tailed significance are colored (grey: *p* > 0.05; cyan blue: *p* < 0.05; dark blue: *p* < 0.01).

#### Age

The observed improvement of including WMH (when adding lesion and bilateral or only right hemispheric WMH) in predicting spatial neglect might be simply an indirect effect of age, due to the known positive correlation between age and WMH extent. We thus asked whether a model using age instead of WMH data yields a similar prediction accuracy. For this purpose, we concatenated the lesion matrix and the vector of normalized age values as we did with the WMH scale ratings above. The averaged model that used lesion map and age as features revealed a prediction accuracy of *R²* = 0.324. Thus, contrary to the feature bilateral/right WMH map, appending the feature age resulted in a less accurate model performance compared to the base model. Further, we calculated a model that attached age as a feature to the model with the best performance so far (i.e., lesion map and right hemispheric WMH map added). We yielded a model fit of *R²* = 0.385, which is again less accurate than the equivalent model without age as a feature. The results indicate that WMH extent is a better predictor than age.

## Discussion

In the present study, we implemented predictive modelling using support vector regression to clarify whether the extent of white matter hyperintensities in addition to voxel-based stroke lesion information is predictive for severity of spatial neglect in acute stroke patients. We compared different measures of WMH extent, namely rating-based measures assessed via ordinal visual rating scales and voxel-based (volumetric, topographic) measures quantified via a semi-automatic demarcation procedure. The combination of volumetric lesion maps and WMH maps yielded improved accuracies compared to the stroke lesion volume-map alone. This finding supports observations by Bahrainwala and colleagues (2014) who observed an association between WMH severity and the probability of having any and also more severe spatial neglect after an acute right hemispheric stroke. The finding is also in line with a study by Meier and colleagues (2020), demonstrating an improved accuracy in predicting cognitive deficits when including WMH extent compared to a base model without WMH information. We observed no increase in model performance when implementing age as an additional feature, although WMH extent and age were moderately correlated. Hence, our analyses revealed that this improvement in predicting neglect severity by including WMH extent information is no indirect effect of age.

The severity of behavioral outcome and lesion size are correlated for many poststroke deficits (Sperber, 2022), and specifically for spatial neglect (Karnath et al., 2004). One could thus argue that the increase of accuracy in predicting spatial neglect by adding the WMH extent to the stroke lesion size is a trivial observation. However, the present findings demonstrate that this assumption does not hold. We observed that the prediction accuracy significantly improved when adding the voxel-based WMH map to the lesion map. In contrast, the CHS-scale did not achieve an improved model performance. But since the CHS-rating correlated strongly with the total WMH volume (and is therefore comparable to it), our findings indicate that the WMH volume per se did not generally benefit the model. Rather the combined information of volume and topography by means of a voxel-based map improved the prediction accuracy. The present results thus demonstrated that the subject-specific joint information of WMH volume and location is essential for an improved prediction of neglect severity. Further, our findings suggest that the voxel-based WMH map is most relevant when treating it as part of the brain lesion itself via direct addition to the lesion map, and less relevant when treating both maps separately. In other words, the anatomical information was most predictive when we treated WMH and the acute stroke lesion as if they constituted the same entity. This finding implies that WMH, although different in etiology, size, and chronicity compared to an acute major ischemic event, might be similar to a stroke lesion in regards to their impact on brain function.

Moreover, our study revealed that prediction via SVR using voxel-based WMH maps as topographic measures outperformed prediction using ordinal visual scale ratings that Bahrainwala and colleagues (2014) used in their study. A volumetrically quantified WMH map not only improved predictability by up to 5.1% (when directly compared to scale ratings), but also provides a potentially more reliable measure. While our study detected WMH by a semi-automated algorithm, scale ratings – such as the CHS-scale (Manolio et al., 1994) or the scale by Fazekas (Fazekas et al., 1987) – are dependent on the visual evaluation of the individual rater and his or her interpretation of the specific scale levels. This circumstance is documented by the rather low consensus in terms of interrater reliability, especially for the Fazekas DS-WMH score in our study. A recent investigation that used both CHS and Fazekas scales reported comparable observations: interrater reliability was moderate for the CHS scale and lower for the Fazekas subscales (Meier et al., 2020). A further aspect arguing in favour of (semi-)automated algorithms for the detection of WMH might be that they allow to detect subtle WMH that would have been overlooked by purely visual inspection but might already constitute diffuse neuropathological changes (Grafton et al., 1991; Schmidt et al., 2011). Rating-based WMH measures, on the other hand, are not suited for detecting potentially relevant nuances and subtle patterns of brain damage.

### Hemisphere effects

WMH of presumed vascular origin are mostly symmetrically in both hemispheres. Interestingly, our study revealed that only ipsilesional, right hemispheric (and bilateral) WMH maps, but not contralesional, left hemispheric WMH maps, were significantly important for the accuracy of predicting spatial neglect. In line with this view, Chen and co-workers (2020) reported that affected white matter fibers of especially the right hemisphere were linked to cognitive impairments in patients with WMH. What might our finding mean that right hemispheric WMH are more relevant than left hemispheric WMH? It is well known that the right hemisphere is dominant in spatial attention (e.g., Becker & Karnath, 2007) and damage to right hemispheric cortical and white matter areas underlie spatial neglect (Karnath et al., 2004; Karnath & Rorden, 2012; Vaessen et al., 2016; Wiesen et al., 2019). An alteration of the right hemispheric white matter microstructure that occurred already prior to stroke might lead to partial disconnections between neglect-related areas of the right hemisphere. The brain would henceforth be more vulnerable to develop attentional deficits poststroke as implied by the concept of ‘brain reserve’ (e.g., Umarova et al., 2021; for a review on brain reserve and spatial neglect, see Umarova, 2017). In a nutshell, the healthier and more sophisticated a brain is, the less it will be affected by events such as stroke, because the brain can tolerate more damage due to a greater reserve. The present study therefore contributes to the discussion on the impact of brain reserve on poststroke cognition and elucidates that WMH are critical for one of the most debilitating neuropsychological disorders after a right hemispheric stroke. In summary, the underlying mechanisms of these findings might contribute to the observation that severe ipsilesional, right hemispheric WMH are relevant for predicting deficits in visual attention after stroke.

### Periventricular WMH (PV-WMH) versus deep subcortical WMH (DS-WMH)

Recent studies proposed the concept of ‘white matter bottlenecks’ whose associated areas appear to be critical in the development of cognitive impairment. These bottleneck regions correspond with the location of PV-WMH and were found to be predictive for language outcome poststroke (Griffis et al., 2017). This supports our conclusion that PV-WMH play a greater role than DS-WMH in predicting neglect severity. In line with this, previous research has reported that PV-WMH were related to deficits in cognition – as, e.g., visuospatial processing in Parkinson’s disease (Huang et al., 2020) and language function in primary progressive aphasia (Meier et al., 2020). Compared to DS-WMH, a higher relative risk of having dementia was also attributed to PV-WMH (Georgakis et al., 2019). However, opposite results were obtained by Kamakura and colleagues (2017) who observed a stronger contribution of DS-WMH compared to PV-WMH in the relationship of severe WMH and worse subacute neglect.

Furthermore, we found only a weak correlation between PV-WMH and DS-WMH. This supports the previous assumption that both WMH variants might have separate origins (Armstrong et al., 2020; Wen et al., 2009) and argues against the suggestion of a common cause (DeCarli et al., 2005; Meier et al., 2020). We speculate that these discrepancies in the literature result not only from dissimilar cohorts but also from the various approaches to assessing WMH extent and the associated comparability issues as already mentioned in the meta-analysis by Armstrong and colleagues (2020); especially, visual rating scales causing disagreements in the interpretation of the severity levels and in the scoring of WMH extent (see above). In future research, the demarcation of WMH volumes with (semi-)automated algorithms should have a priority.

### Extent of WMH and chronic neglect

There is first evidence for the association of WMH extent and recovery from neglect. Hawe and colleagues (2018) reported a significant relation between chronic neglect performance and the calculated overall volumes of stroke lesion and WMH, whereas the volumes of lesion and of WMH alone were not associated with neglect recovery. Their finding corresponds well with our observation that the combination of stroke lesion and WMH volume-maps is a better predictor than one of those maps alone for neglect severity in acute stroke patients. Hawe and colleagues (2018) concluded that the stroke lesion alone did not cause chronic neglect as severe as when extensive WMH were combined with it. It is possible that a pre-damaged brain due to WMH formation might be more vulnerable to develop attentional deficits following stroke, maybe due to white matter disconnections. Further, Kamakura and colleagues (2017) found in a case series that less severe WMH were related to less severe neglect in the subacute phase; the degree of WMH extent differed between patients who had or had not recovered from neglect.

### Conclusions

The present study demonstrated that voxel-based WMH information affect the severity of spatial neglect in the acute phase of stroke. Our findings argue in favor of using combined maps of stroke lesion and WMH extent in predictive modelling. These maps might also lead to more accurate results in lesion-behavior mapping. Results further indicate that the combination of precise voxel-based maps of both stroke lesion and ipsilesional, right hemispheric WMH yields an improved accuracy in predicting spatial neglect severity compared to the acute stroke lesion map alone. Since our models with precise volumetric WMH maps as auxiliary predictors resulted in better accuracies than ordinally scaled visual ratings (i.e., CHS-scale and the scale by Fazekas), we recommend measuring WMH extent topographically by means of a voxel-based map. Further studies are needed that apply the approach of the present research to investigate if the voxel-based WMH map results also in a more accurate prediction of other cognitive deficits following stroke. If so, the methodological approach applied and associated results have great potential to become clinically relevant for rehabilitation as WMH appears to constitute a critical imaging biomarker for (poststroke) cognitive impairment.

## Data Availability

Data containing patient information cannot be made publicly available due to data protection restrictions by the local ethics commission. Remaining data and custom scripts are openly available in Mendeley Data (http://dx.doi.org/10.17632/c8n42jz525.1).

https://data.mendeley.com/datasets/c8n42jz525/1

## Acknowledgments

This work was supported by the Deutsche Forschungsgemeinschaft (KA 1258/23-1) and National Institutes of Health (P50DC014664). We would like to thank Vanessa Kasties for advice and discussion on machine learning methods.

## CredIt author contribution statement

*Lisa Röhrig*: Conceptualization, Formal analysis, Investigation, Methodology, Visualization, Writing – original draft;

*Christoph Sperber*: Methodology, Writing – review & editing;

*Leonardo Bonilha*: Conceptualization, Writing – review & editing;

*Christopher Rorden*: Conceptualization, Writing – review & editing;

*Hans-Otto Karnath*: Conceptualization, Data curation, Funding acquisition, Resources, Supervision, Writing – review & editing.

## Supplemental Figures

**Figure S1.**
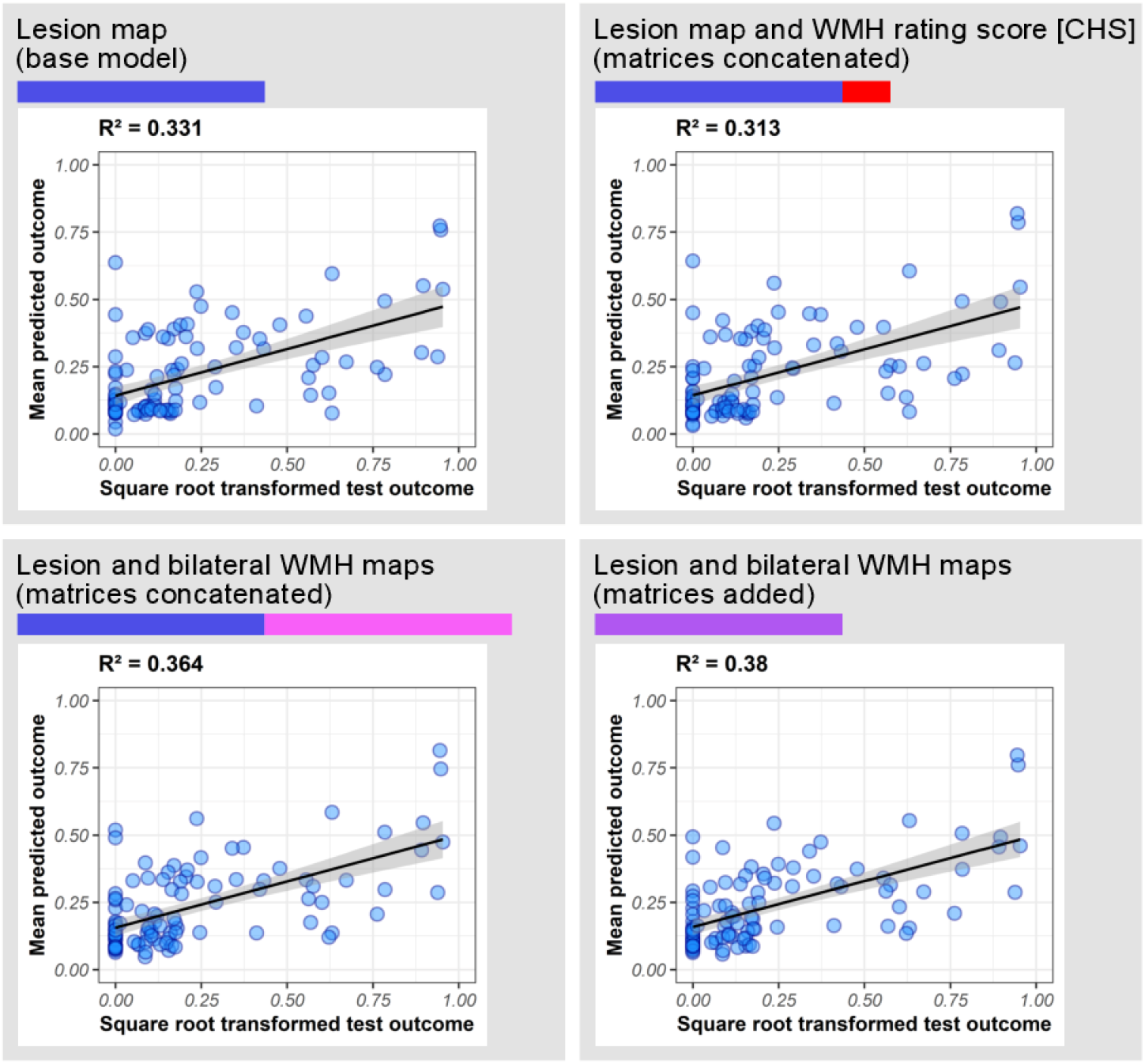
Model fits across model versions. Performances of the cross-validated and averaged models are displayed for prediction of neglect severity (CoC score). The different feature matrices are visualized as colored bars as in Figure 2 (main article). *Coefficient of Determination* (*R²*) is reported as prediction accuracy. Scatter plots show the relationship of the test outcomes and the predictions averaged across 10 model repetitions; a linear model line is depicted with its regarding 95%-confidence interval (grey).

**Figure S2.**
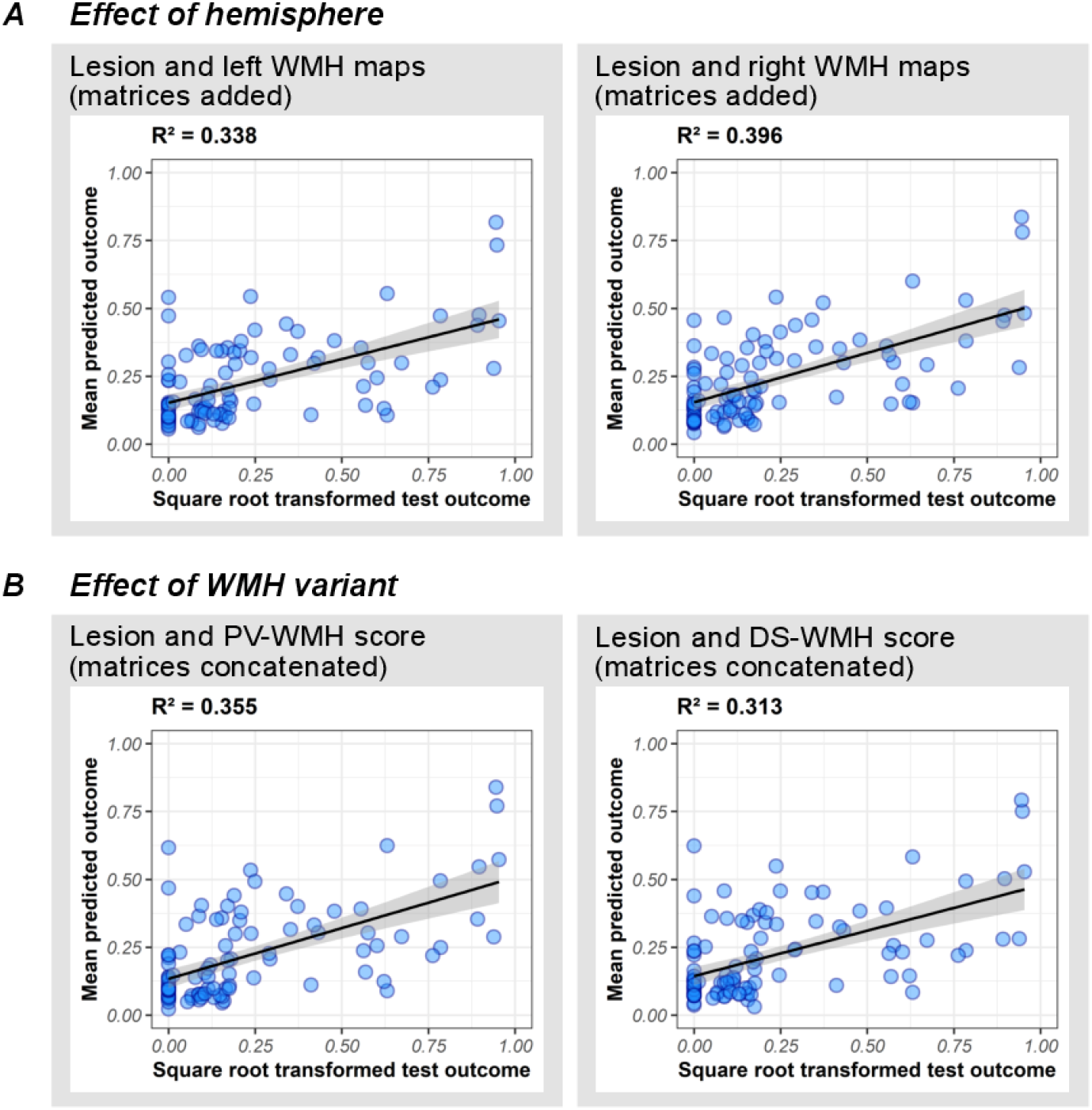
Effect of white matter hyperintensities (WMH) localization on model fits. Illustrated are the effects of (**A**) hemisphere (left versus right hemispheric WMH) and (**B**) WMH variant (periventricular WMH [PV-WMH] versus deep subcortical WMH [DS-WMH]) on the model fits. For the effect of hemisphere, lesion and unilateral WMH maps were added. For the effect of WMH variant, lesion map and Fazekas rating scores were concatenated. Performances of the cross-validated and averaged models are displayed for the prediction of neglect severity (CoC score). *Coefficient of Determination* (*R²*) is reported as prediction accuracy. Scatter plots show the relationship of the test outcomes and the predictions averaged across 10 model repetitions; a linear model line is depicted with its regarding 95%-confidence interval (grey).

